# Hs-CRP is associated with Heart Failure Hospitalization in Patients with MAFLD and Normal LVEF Undergoing Coronary Angiography

**DOI:** 10.1101/2023.10.06.23296684

**Authors:** Xiao-Dong Zhou, Qin-Fen Chen, Giovanni Targher, Christopher D. Byrne, Michael D. Shapiro, Na Tian, Tie Xiao, Ki-Chul Sung, Gregory Y. H. Lip, Ming-Hua Zheng

## Abstract

**Background:** Systemic chronic inflammation plays a role in the pathophysiology of both heart failure with preserved ejection fraction (HFpEF) and metabolic dysfunction-associated fatty liver disease (MAFLD).

**Aim:** This study aimed to investigate whether serum high-sensitivity C-reactive protein (hs-CRP) levels were associated with the future risk of heart failure (HF) hospitalization in patients with MAFLD and a normal left ventricular ejection fraction (LVEF).

**Methods:** The study enrolled consecutive individuals with MAFLD and normal LVEF who underwent coronary angiography for suspected coronary heart disease. The study population was subdivided into non-HF, pre-HFpEF, and HFpEF groups at baseline. The study outcome was the first hospitalization for HF.

**Results:** In 10,019 middle-aged individuals (mean age 63.3±10.6 years; 38.5% female), the prevalence rates of HFpEF and pre-HFpEF were 34.2% and 34.5%, with a median serum hs-CRP level of 4.5 mg/L (IQR: 1.9-10 mg/L) and 5.0 mg/L (IQR: 2.1-10.1 mg/L), respectively. Serum hs-CRP levels were significantly higher in the pre-HFpEF and HFpEF groups than in the non-HF group. HF hospitalizations occurred in 1942 (19.4%) patients over a median of 3.2 years, with rates of 3.7% in non-HF, 20.8% in pre-HFpEF, and 32.1% in HFpEF, respectively. Cox regression analyses showed that patients in the highest hs-CRP level quartile had a ∼4.5-fold increased risk of being hospitalized for HF compared to those in the lowest hs-CRP level quartile (adjusted-Hazard Ratio 4.42, 95% CI 3.72-5.25).

**Conclusions:** There was a high prevalence of baseline pre-HFpEF and HFpEF in subjects with MAFLD. There was an increased risk of HF hospitalization in those with elevated hs-CRP levels.

## Introduction

Metabolic dysfunction-associated fatty liver disease (MAFLD), formerly named non-alcoholic fatty liver disease, is a highly prevalent metabolic liver condition worldwide, affecting up to nearly 30% of the general adult population.^1–5^ Recent cohort studies suggested that patients with MAFLD have an increased risk of developing new-onset heart failure (HF), especially HF with preserved ejection fraction (HFpEF).^6–8^ A comprehensive meta-analysis of longitudinal cohort studies (including ∼11 million middle-aged individuals from different countries) showed that MAFLD was associated with a 1.5-fold increased risk of new-onset HF over a median of 10 years.^9^

Despite a normal left ventricular ejection fraction (LVEF), HFpEF is a common chronic cardiac condition globally, where metabolic dysfunction (e.g., obesity and type 2 diabetes) and low-grade chronic inflammation may contribute importantly to its pathogenesis.^10–12^ HFpEF is associated with a substantially higher risk of adverse cardiovascular events and all-cause mortality.^10–12^

Empiric evidence suggests that the unifying link between MAFLD and HFpEF is low-grade chronic inflammation, which may adversely affect cardiomyocyte function.^13–16^ This low-grade inflammatory state is characterized by an increase in various biomarkers in the bloodstream.^17,18^ For example, high-sensitivity C-reactive protein (hs-CRP) is one of the most widely used biomarkers for systemic inflammation, and an increase in hs-CRP is predictive of adverse cardiovascular events, such as myocardial infarction, stroke, and HF.^19–21^ However, to our knowledge, the ability of serum hs-CRP level to predict future HF events in patients with MAFLD and preserved LVEF has not been explored.

The main aims of our study were as follows: (1) to examine the prevalence of HFpEF among patients with MAFLD and suspected coronary artery disease (CAD) undergoing elective coronary angiography; and (2) to evaluate the associations between increased serum hs-CRP levels and the future risk of HF hospitalizations in this patient population.

## Methods

### Study Design

This retrospective longitudinal study enrolled individuals diagnosed with MAFLD and suspected CAD who had undergone conventional echocardiograms at the First Affiliated Hospital of Wenzhou Medical University between January 2009 and February 2023. The inclusion criteria were as follows: (1) aged 18 years or older; (2) diagnosis of MAFLD; (3) presence of LVEF ≥50% on echocardiography; and (4) acceptance to undergo an elective coronary angiography. Patients who did not meet the inclusion criteria (mentioned above), patients who were unable to provide informed consent, those who had had any acute inflammatory condition, and any other organ failure, rheumatological disorder, malignancy, or those lost at follow-up, were excluded from the study (as specified in **Supplementary Figure 1**).

Baseline data for all patients were collected retrospectively through electronic medical records, which provided various details such as medical history, demographic variables, clinical and laboratory data, current use of medications, liver ultrasound results, echocardiography evaluation findings, and subsequent follow-up data.

The study was conducted in compliance with the Declaration of Helsinki, and the ethics committee of the First Affiliated Hospital of Wenzhou Medical University approved the study protocol with a waiver for informed consent due to the infeasibility of obtaining informed consent given the study’s retrospective design.The data that support the findings of this study are available from the first author upon reasonable request.

### MAFLD Diagnosis

In all patients, MAFLD was diagnosed by the presence of hepatic steatosis on liver ultrasound or blood biomarkers/scores in combination with at least one of the following metabolic risk factors: overweight/obesity, type 2 diabetes, or at least two of the following metabolic abnormalities: 1) waist circumference ≥90/80 cm in men and women; 2) blood pressure ≥130/85 mmHg or specific drug treatment; 3) serum triglycerides ≥150 mg/dl (≥1.70 mmol/L) or specific drug treatment; 4) serum high-density lipoprotein (HDL)-cholesterol <40 mg/dl (<1.0 mmol/L) for men and <50 mg/dl (<1.3 mmol/L) for women or specific drug treatment; 5) prediabetes, defined as fasting glucose levels between 100 to 125 mg/dl [5.6 to 6.9 mmol/L], or HbA1c levels ranging from 5.7% to 6.4% [39 to 47 mmol/mol]; 6) a Homeostasis Model Assessment (HOMA) score for insulin resistance of ≥2.5; and 7) a plasma hs-CRP level of >2 mg/L.^22,23^ FIB-4 index was calculated as follows: age × aspartate aminotransferase (U/L)/[platelet (10^9^/L) × alanine aminotransferase1/2 (U/L)].^24^ Serum hs-CRP was measured using an immuno-turbidimetry assay in a Beckman Coulter analyzer (AU5800).

### Baseline HF status

The study population was subdivided into the non-HF, pre-HFpEF, and HFpEF groups according to the presence or absence of HF symptoms and impaired cardiac function at baseline. The diagnosis of pre-HFpEF was defined as asymptomatic patients (absence of signs or symptoms of HF) with ‘preserved’ ejection fraction (LVEF ≥50%) who had at least one of the following conditions: evidence of structural heart disease (including left atrial enlargement), and/or diastolic dysfunction, presence of multiple cardiovascular risk factors with elevated levels of natriuretic peptides, or persistently elevated cardiac troponins, in the absence of competing diagnoses.^25,26^ The diagnosis of HFpEF was defined as symptomatic patients with ‘preserved’ ejection fraction (LVEF ≥50%) who had at least one of the following conditions: evidence of structural heart disease (including left atrial enlargement) and/or diastolic dysfunction, multiple cardiovascular risk factors with elevated levels of serum natriuretic peptides, or persistently elevated cardiac troponins, in the absence of competing diagnoses.^25,26^ In contrast to ‘true HFpEF’, the key clinical component of pre-HFpEF was the absence of HF signs and symptoms.

### Coronary Angiography

All study patients underwent elective coronary angiography to quantify the presence of CAD. The reports of coronary angiographies of all patients were meticulously reviewed and categorized in cooperation with the study’s cardiologist, X-D Zhou. Mild CAD was defined as coronary stenoses <50%, moderate CAD as stenoses 50-70%, and severe CAD as having at least one proximal coronary artery with >70% stenosis based on angiography.^27^

### Study Outcomes

Clinical follow-up data were collected from inpatient and outpatient medical records to analyze the clinical study outcome. The length of the follow-up was determined as the time between the MAFLD diagnosis and the first occurrence of either the end of clinical follow-up or the time-to-event endpoints, whichever came first. Patients were followed until April 2023 to examine the clinical outcome for prognostic purposes systematically. The primary outcome of the study was the first hospitalization for HF.

### Statistical Analysis

All statistical analyses were performed using the IBM SPSS software, version 23.0 for Windows. Continuous variables were expressed as means ± SD or medians (interquartile ranges, IQR), and categorical variables as percentages. Statistical comparisons between the study groups were carried out using the unpaired Student’s *t*-test (for normally distributed continuous variables), the Mann-Whitney U test (for non-normally distributed continuous variables), and the chi-squared test (for categorical variables). We performed unadjusted and adjusted Cox proportional hazards models to examine the association between serum hs-CRP levels (stratified by increasing quartiles, from Q1 to Q4) and the risk of HF hospitalization during the follow-up period. The Cox proportional hazards models provided the hazard ratios (HR) and 95% confidence intervals (CI). Furthermore, a Kaplan-Meier survival analysis was also performed to calculate the event-free survival curves, and the log-rank test was used to test the presence of any significant differences between the curves. A statistically significant level was considered as P-value <0.05 (two-tailed).

## Results

### Baseline Characteristics

The final sample for analysis consisted of 10,019 middle-aged Chinese patients (mean age 63.3±10.6 years; 38.5% female) with MAFLD and suspected CAD who underwent elective coronary angiography, after excluding patients who did not meet the study’s inclusion criteria (**Supplementary Figure 1**). At baseline, 3,133 (31.3%) patients had non-HF, 3,427 (34.2%) had pre-HFpEF, and 3,459 (34.5%) had HFpEF, respectively. Detailed baseline characteristics, traditional cardiovascular risk factors, and laboratory parameters of patients stratified by different baseline HF statuses are shown in **Table 1**. Patients with HFpEF were older, had more comorbidities, a more atherogenic risk profile, a greater prevalence of severe coronary stenosis, larger left ventricular end-diastolic diameter, higher FIB-4 score, and lower HSI score compared to the other two patient groups. Serum hs-CRP levels both in the pre-HFpEF group (4.5 mg/L; IQR: 1.9-10 mg/L) and in the HFpEF group (5.0 mg/L; IQR: 2.1-10.1 mg/L) were significantly higher than those in the non-HF group (2.7 mg/L; IQR: 1.1-5.0 mg/L).

**Table 1.**
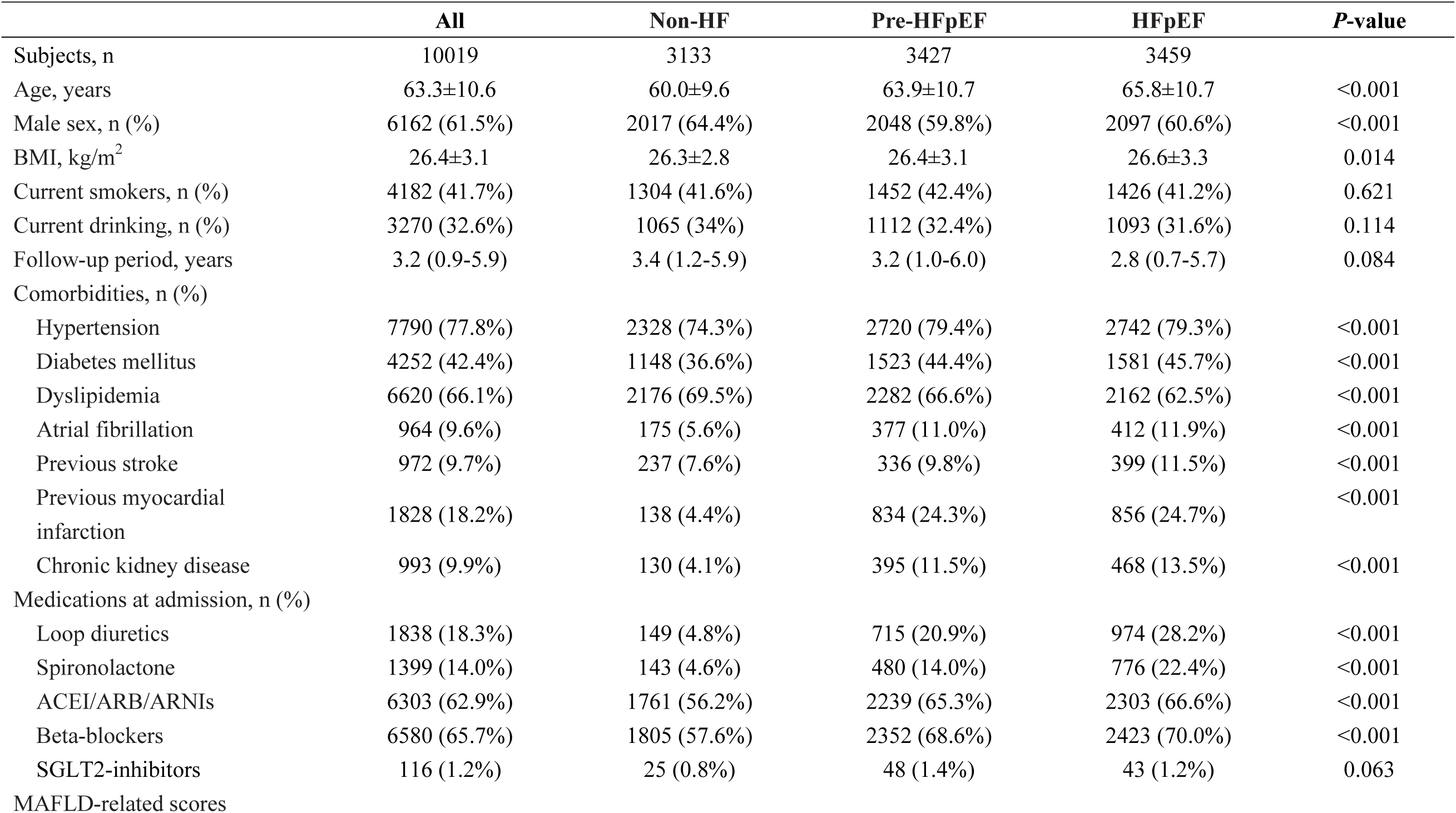
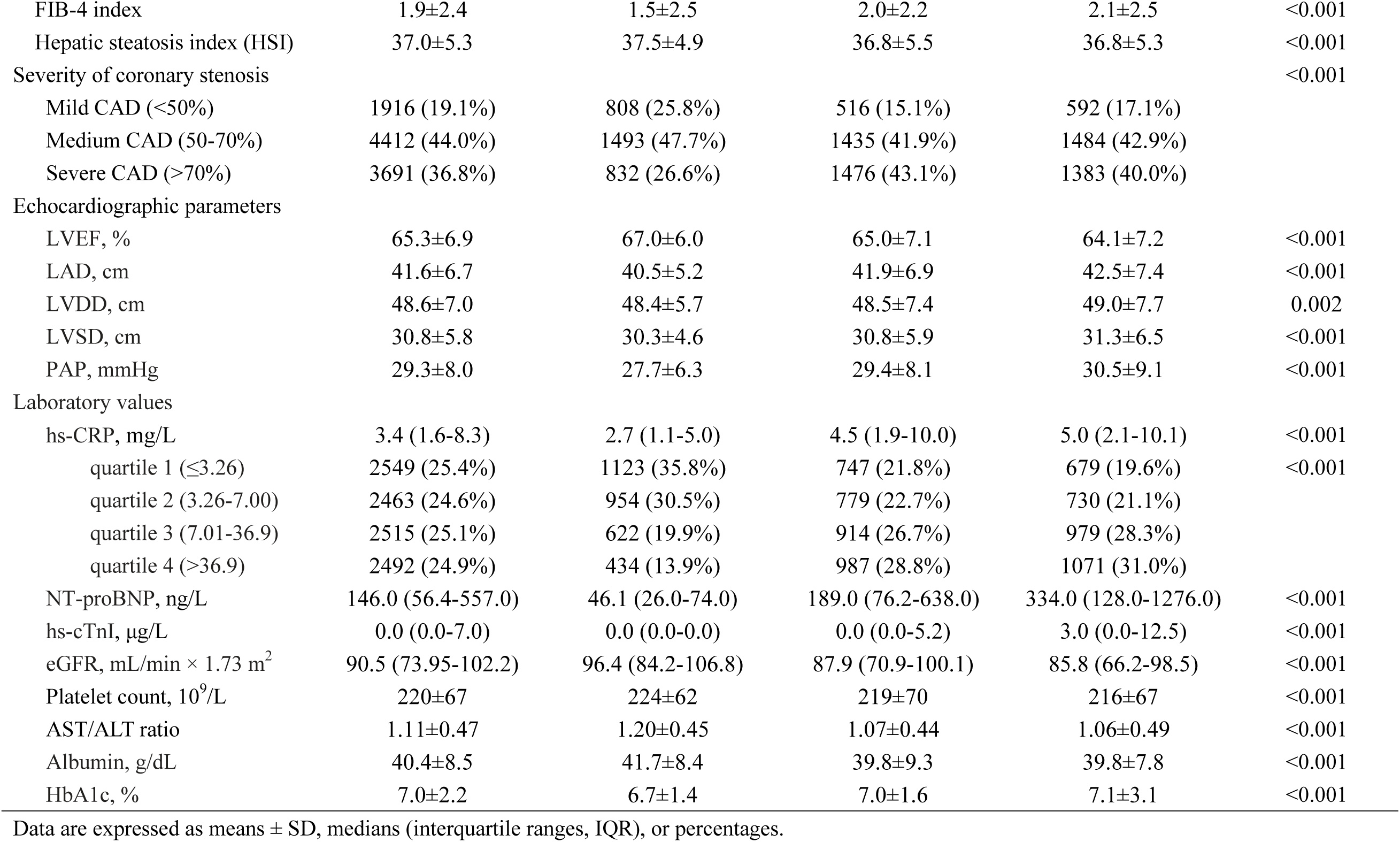
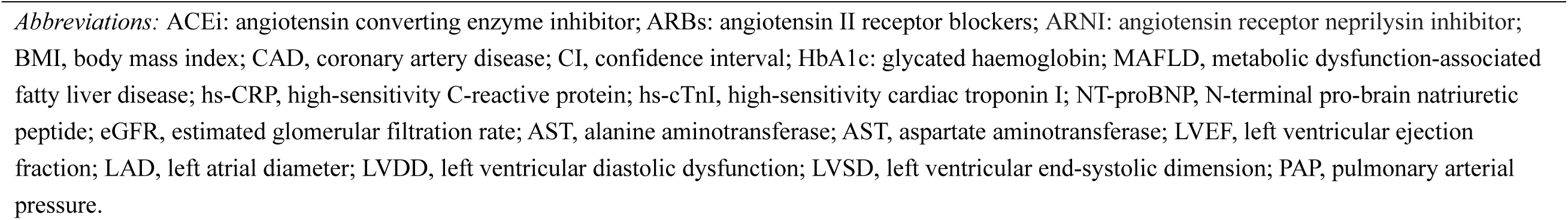
Baseline Clinical and Biochemical Characteristics of Patients with MAFLD and Suspected Coronary Artery Disease Stratified by Baseline Heart Failure Status.

### Pre-HFpEF and HFpEF prevalence, and incident HF hospitalization

As shown in **Figure 1**, about two-thirds of patients had pre-HFpEF or HFpEF and the prevalence rates of these two cardiac conditions increased across quartiles of serum hs-CRP at baseline. During a median follow-up period of 3.2 years (IQR: 0.9-5.9 years), hospitalizations for HF occurred in 1942 (19.4%) patients, with an incidence rate of 6.1 events per 100 person-years. As also shown in the figure (panel C), patients with HFpEF or pre-HFpEF at baseline were more likely to be hospitalized for HF than those in the non-HF group.

**Figure 1.**
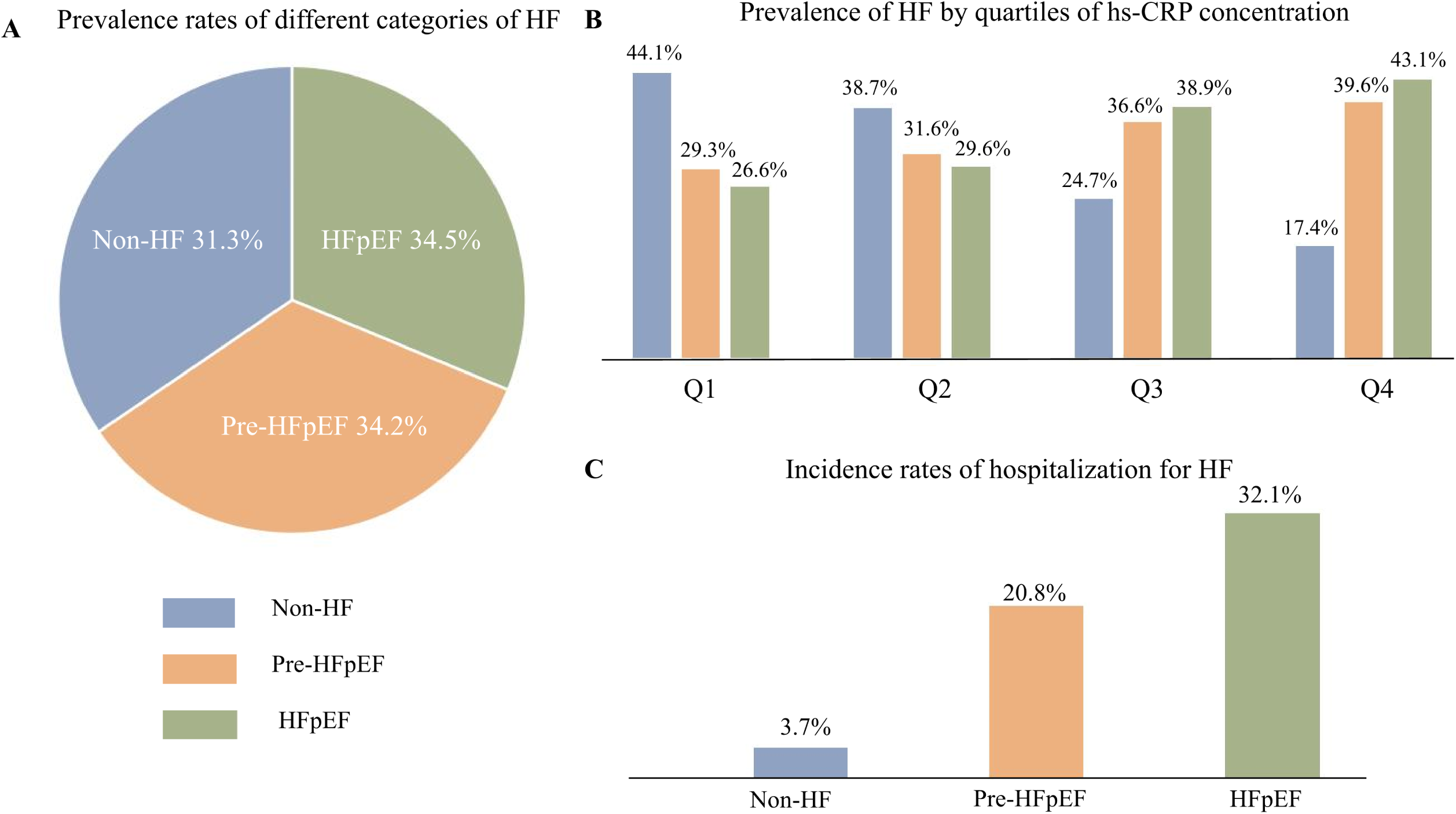
Prevalence rates of different categories of HF (i.e., non-HF, preHFpEF, and HFpEF) in the whole cohort (A) and patients stratified by quartiles (Q1 to Q4) of serum hs-CRP concentrations (B). Incidence rates of hospitalization for HF according to different HF categories at baseline (C).

### Hs-CRP and risk of incident HF hospitalization

As shown in **Table 2**, patients in the highest baseline quartile of hs-CRP levels had a markedly higher risk of HF hospitalization compared to those in the lowest hs-CRP quartile (unadjusted HR 6.937, 95% 5.857-8.215). Increased serum hs-CRP levels were significantly associated with a higher risk of HF hospitalization (HR 4.421, 95% 3.720-5.254), even after adjustment for age, sex, smoking history, alcohol intake, BMI, hypertension, diabetes, dyslipidemia, atrial fibrillation, previous stroke, previous myocardial infarction, chronic kidney disease, and current use of loop diuretics, spironolactone, ACEI/ARB/ARNIs or beta-blockers. A Kaplan-Meier survival analysis showed a significant incremental increase in the risk of HF hospitalization across serum hs-CRP quartiles (P <0.001 by the log-rank test, **Figure 2**).

**Table 2.**
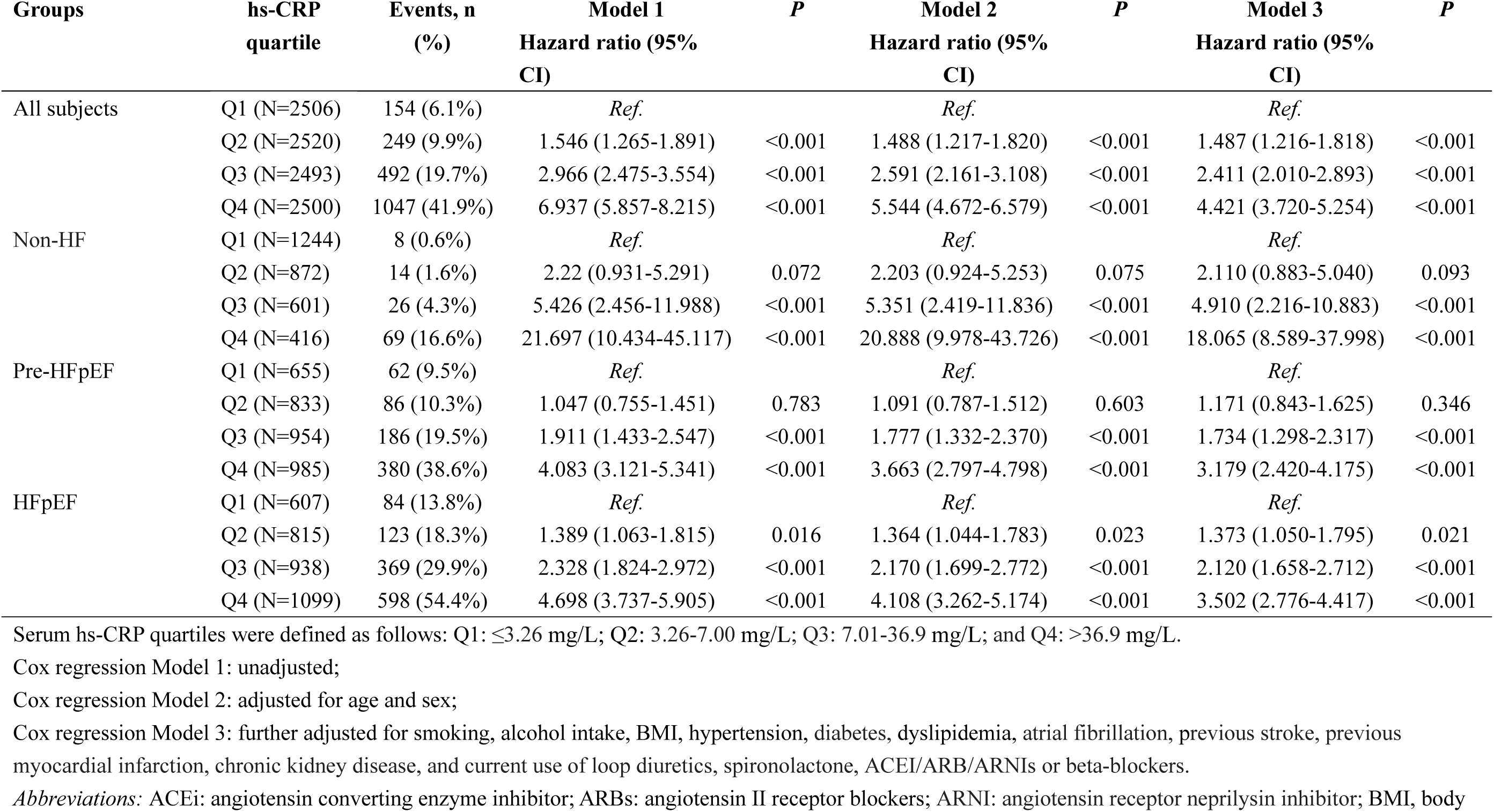
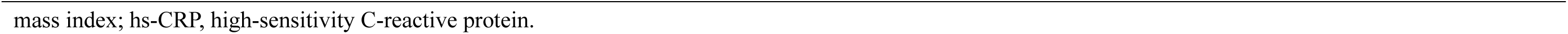
Associations between hs-CRP Concentration Quartiles and the Risk of Heart Failure Hospitalization in Patients with Different Heart Failure Status at baseline.

**Figure 2.**
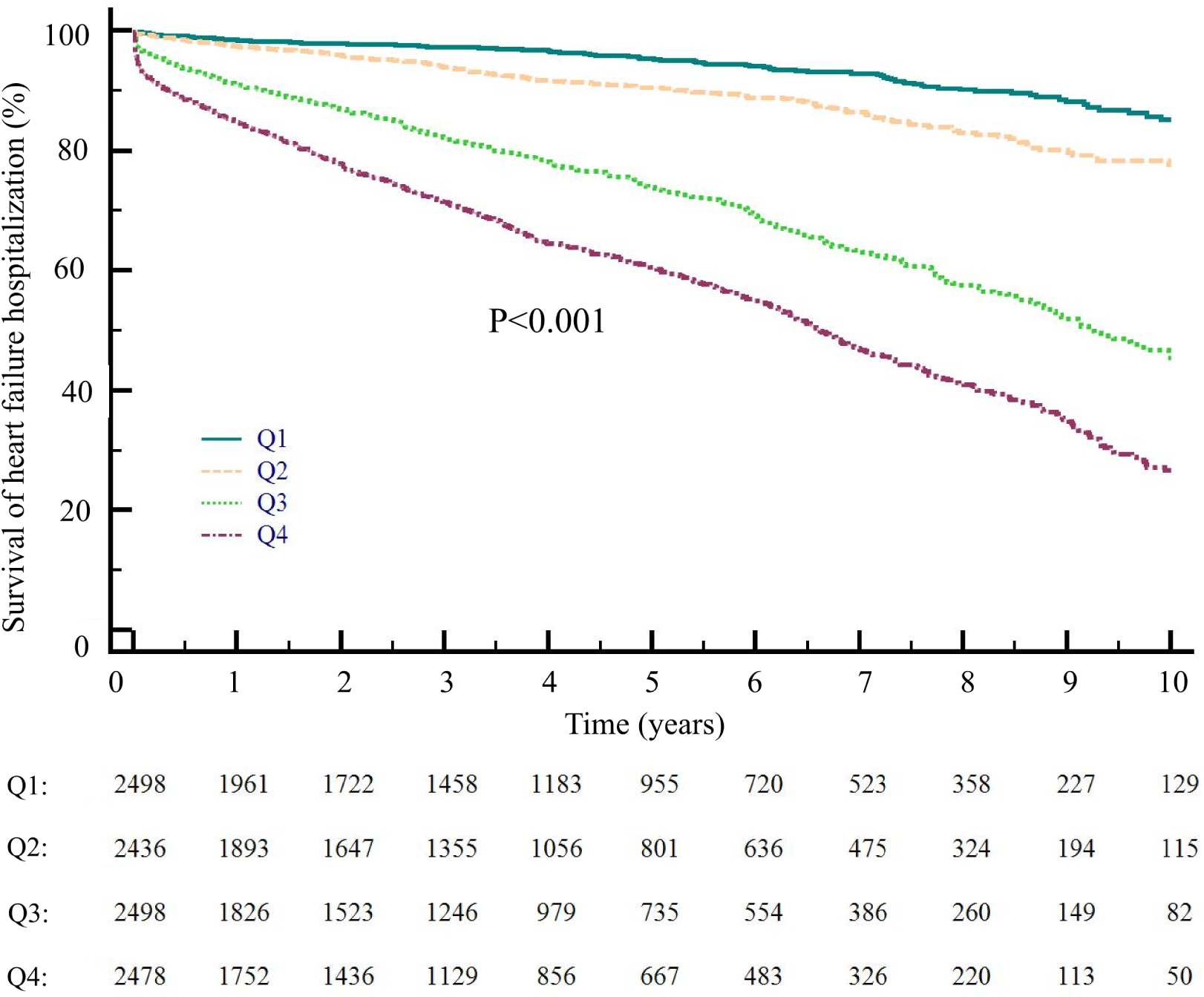
Kaplan-Meier event-free survival curve of the risk for HF hospitalizations in the whole cohort of patients stratified by serum hs-CRP quartiles. P-values were tested by log-rank test. Under the x-axis are reported the number of subjects in each quartile at each time.

### Hs-CRP and increased risk of HF hospitalization in subgroups

We performed subgroup analyses to examine the significant associations between serum hs-CRP quartiles and the risk of HF hospitalization. This risk remained statistically significant even after adjusting for potential confounders, i.e., regardless of the HF status (**Table 2** and **Figure 3**), the severity of coronary stenoses (**Supplementary Table 1** and **Figure 3**), or FIB-4 score at baseline (**Supplementary Table 2** and **Figure 4**).

**Figure 3.**
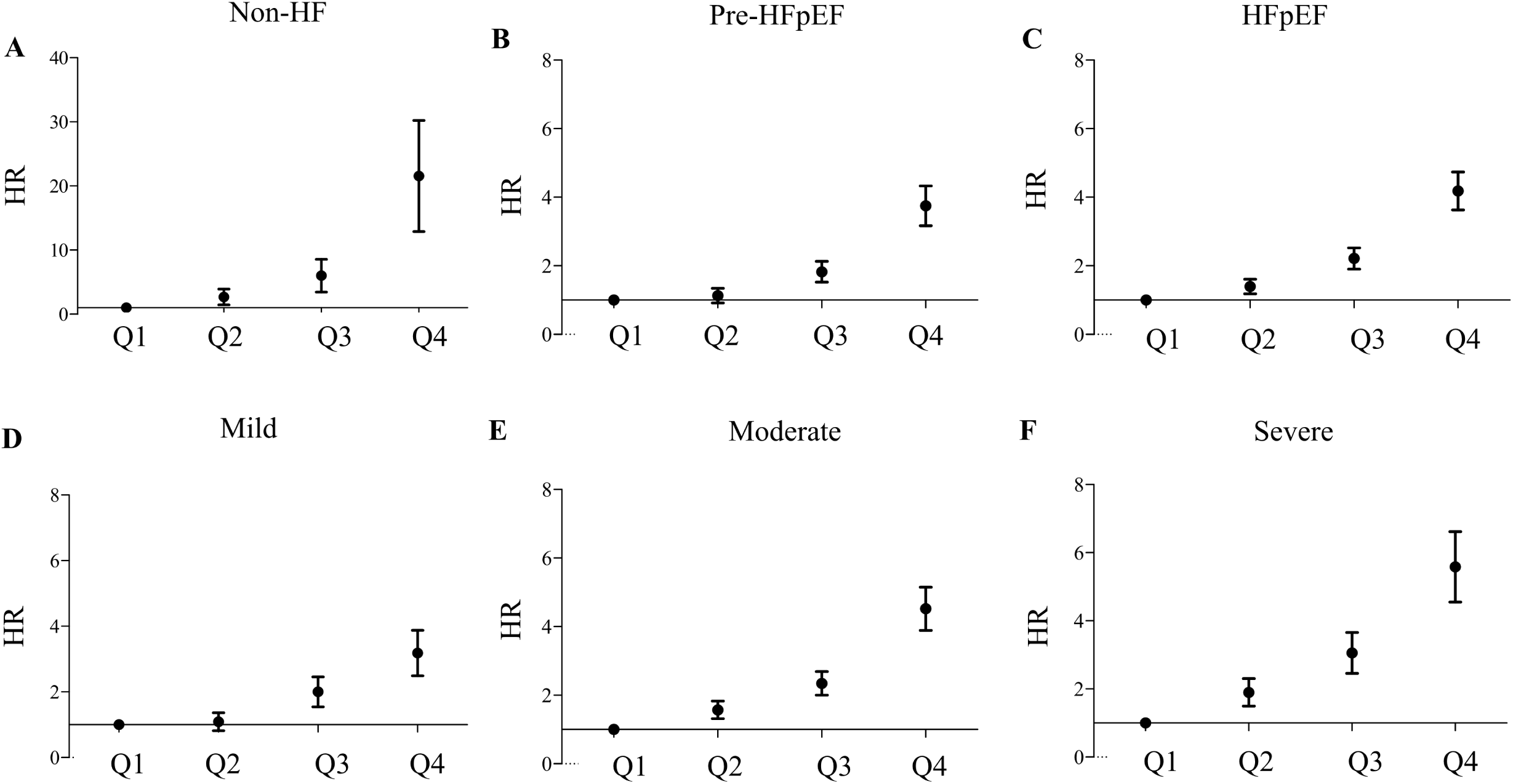
Hazard ratios (HR) and 95% confidence intervals for HF hospitalization in patients stratified by different status of HF and severity of coronary stenosis at baseline: (A) Non-HF; (B) Pre-HFpEF; (C) HFpEF; (D) Mild stenosis; (E) Moderate stenosis; and (F) Severe stenosis.

**Figure 4.**
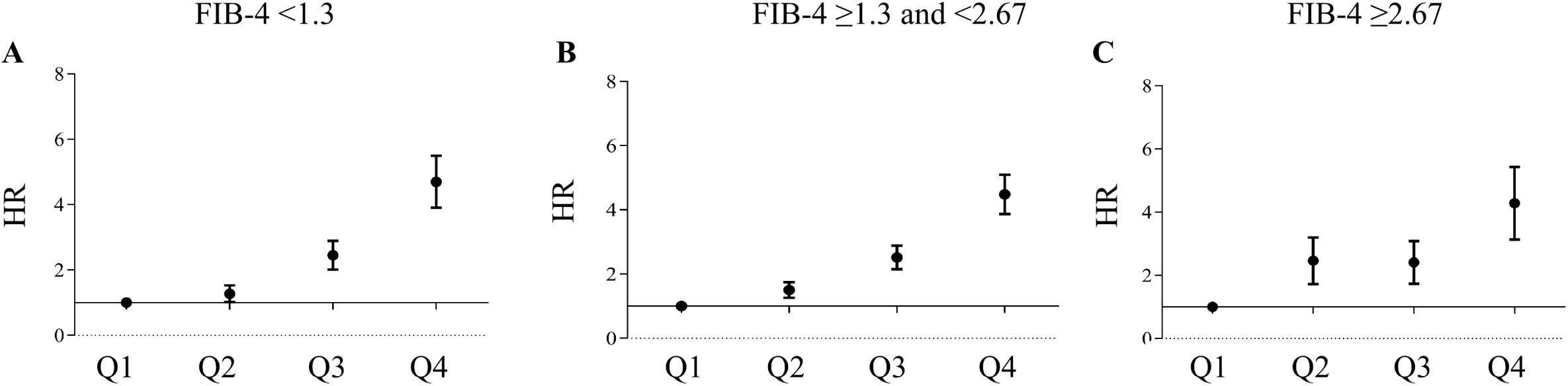
Hazard ratios (HR) and 95% confidence intervals for HF hospitalization in patients stratified by MAFLD-related scores: (A) FIB-4 <1.3; (B) FIB-4 ≥1.3 and < 2.67; (C) FIB-4 ≥2.67.

## Discussion

The key findings from this analysis are summarized as follows: (1) pre-HFpEF and HFpEF are two highly prevalent cardiac conditions affecting up to nearly two-thirds of this patient population with MAFLD; (2) patients with pre-HFpEF or HFpEF are at higher risk of being hospitalized for HF than the non-HF patient group, with incidence rates of 3.7% in non-HF, 20.8% in pre-HFpEF, and 32.1% in HFpEF, respectively; (3) serum hs-CRP levels are increased in patients with pre-HFpEF or HFpEF; and increased hs-CRP levels predicted the future risk of hospitalization for HF, regardless of the different HF status and the severity of coronary stenosis at baseline.

While serum hs-CRP levels are closely associated with an elevated risk of adverse cardiac events in individuals with cardiometabolic disease, there is limited data that specifically evaluate the connections between serum hs-CRP and HFpEF in patients with MAFLD. Our study provides novel data from a large cohort of MAFLD patients with suspected CAD to address this question.

### Prevalence of HFpEF in MAFLD

Patients with MAFLD often have multiple cardiometabolic disorders leading to myocardial remodeling and diastolic dysfunction over time.^28–30^ However, these individuals are more likely to develop HFpEF than patients with HF with reduced LVEF (HFrEF).^31,32^ Hence, understanding the prevalence of pre-HFpEF and HFpEF among patients with MAFLD is clinically important for promptly identifying individuals at higher risk of developing HF and who may benefit from targeted pharmacotherapies to reduce their HF risk.

In the present large study, a significant proportion of our individuals with MAFLD and normal LVEF had pre-HFpEF or HFpEF (about 34% for every condition). Moreover, the overall rates of HF hospitalization we observed in our study were nearly 5-8 times greater in the pre-HFpEF and HFpEF groups than in the non-HF group, with rates of 20.8% in pre-HFpEF vs. 32.1% in HFpEF vs. 3.7% in non-HF.

### Chronic inflammation may link MAFLD to HFpEF

Low-grade chronic inflammation is a common mechanism that may pathophysiologically link MAFLD to the development and progression of HFpEF.^13,33,34^ MAFLD, especially in its more advanced histological forms, may exert adverse effects mainly through the systemic release of multiple pro-inflammatory, pro-oxidant, and pro-fibrotic mediators, thus contributing to the development of various extrahepatic complications, including functional and structural cardiac abnormalities that can lead to new-onset HFpEF.^35–38^

### Hs-CRP levels, pre-HFpEF or HFpEF and the future risks of HF hospitalization

The findings of our study highlight the importance of measuring serum hs-CRP levels in patients with MAFLD and suspected CAD and represent an essential consideration for hepatologists when assessing the future risk of HF hospitalization in this patient population. Hepatologists may overlook hs-CRP measurements when MAFLD presents with preserved LVEF and no obvious signs and symptoms of HF.

In our study, we found that compared to those with the lowest serum hs-CRP levels, patients with increased hs-CRP levels not only had significantly higher prevalence rates of pre-HFpEF and HFpEF but also had higher incidence rates of HF hospitalization over a mean period of 3.2 years, irrespective of the severity of coronary stenosis or different HF status at baseline. Thus, serum hs-CRP may be a biomarker for predicting the future risk of HF hospitalization in patients with MAFLD. The present findings also suggest that hepatologists need to pay greater attention to the potential risk of HF in patients with MAFLD and normal LVEF.

### Limitations

The current study has some important limitations. First, we conducted the research retrospectively at a single academic center, which may have resulted in selection bias. Second, we acknowledge that the study patients referred for elective coronary angiography may have experienced referral bias, leading to an increased risk of having HF among people suspected of CAD. Third, the length of follow-up was relatively short. Finally, we recognize that using electronic medical records may have led to an underestimation of HF hospitalization rates since these records may not have captured instances where patients were admitted to hospitals outside our institution.

## Conclusions

Among Chinese middle-aged individuals with MAFLD and suspected CAD undergoing elective coronary angiography, there was a high prevalence of baseline pre-HFpEF and HFpEF in subjects with MAFLD. There was an increased risk of HF hospitalization in those with elevated hs-CRP levels.

## Authorship contribution statement

**Xiao-Dong Zhou**: Conceptualization, Formal analysis, Investigation, Data curation, Writing - original draft, Visualization. **Qin-Fen Chen**: Conceptualization, Formal analysis, Investigation, Data curation, Writing - original draft, Visualization. **Giovanni Targher**: Investigation. **Christopher D. Byrne**: Investigation. **Michael D. Shapiro**: Investigation. **Na Tian**: Investigation.**Tie Xiao**: Investigation. **Ki-Chul Sung**: Investigation. **Gregory Y. H. Lip:** Investigation. **Ming-Hua Zheng**: Conceptualization, Investigation, Supervision, Project administration, Funding acquisition, Writing - review & editing.

## Data Availability

The data that support the findings of this study are available from the first author upon reasonable request.

## Tables

**Supplementary Table 1.** Associations between hs-CRP Concentration Quartiles and the Risk of Heart Failure Hospitalization in Patients with Different Severity of Coronary Stenosis at baseline.

**Supplementary Table 2.** Associations between hs-CRP Concentration Quartiles and the Risk of Heart Failure in Patients with MAFLD and Different Categories of MAFLD-related Score.

## Figure Legends

**Supplementary Figure 1.** Flowchart of the study design.

